# An alternative workflow for molecular detection of SARS-CoV-2 - escape from the NA extraction kit-shortage, Copenhagen, Denmark, March 2020

**DOI:** 10.1101/2020.03.27.20044495

**Authors:** Anna S. Fomsgaard, Maiken W. Rosenstierne

**Affiliations:** Department of Virus and Microbiological Special Diagnostics, Statens Serum Institut, Copenhagen, Denmark

**Keywords:** SARS-CoV-2, COVID-19, molecular diagnostics, RT-qPCR, diagnostics

## Abstract

The World Health Organisation has declared a pandemic caused by the newly discovered SARS-CoV-2. Due to growing demand for reagents used for SARS-CoV-2 RNA extraction for subsequent molecular diagnostics, there is a worldwide risk of kit- and/or reagent-shortages for extraction. With a detection sensitivity of 97.4% (95% CI=86.2-99.9%), we describe a simple, fast, alternative workflow for molecular detection of SARS-CoV-2, where samples are simply heat-processed for 5 minutes at 98°C prior to the RT-qPCR reaction.

---

Coronavirus disease (COVID-19) caused by the novel severe acute respiratory syndrome coronavirus-2 (SARS-CoV-2), was first noticed in Wuhan, China in December 2019 and then spread across the globe in few months [1]. Some of the largest manufacturers of reagents for molecular diagnostics, like Qiagen and Roche, are struggling to keep up the global demand for RNA extraction reagents [2]. As we are facing a massive global increase in COVID-19 worldwide, shortage in NA extraction kits, will result in a future where patients cannot be diagnosed for this and other viruses. Here, we describe a new simplified workflow for molecular detection of SARS-CoV-2, without NA extraction, which could serve as an alternative in diagnostic laboratories to overcome chemical based kit-shortage.

## Direct approach for molecular detection of SARS-CoV-2

NA purification prior to PCR/RT-PCR is the golden standard for molecular diagnostics. The MagNa Pure 96 system (Roche Molecular Biochemicals, Indianapolis, Ind.) is a widely used system for high-throughput NA purification in many public health institutions worldwide [3]. However, with Roche’s announcement of emerging kit-shortage or bottlenecks [2], we wanted to investigate if real-time RT-PCR (RT-qPCR) analysis could be performed with minimal pretreatment on patients (n=89) oropharyngeal swaps, which are the most common sample type collected from patients suspected of COVID-19 in several countries, including Denmark.

Three simplified approaches without NA purification were performed before RT-qPCR for SARS-CoV-2: 1) *Direct*: the saline/transport solution from the throat-swap, 2) *PBS diluted*: the saline/transport solution was further diluted 1:1 with phosphate-buffered saline (PBS), and 3) *Heat-processed*: We tested four different heat-processes: an aliquote of the saline/transport solution was heat-processed for a) 5 min. at 95°C; b) 10 min. at 95°C; c) 5 min. at 98°C; and d) 10 min. at 98°C, respectively. All heat-processed clinical samples were cooled for 2 min. at 4°C before being used in the RT-qPCR reaction. Two SARS-CoV-2 RT-qPCR assays were used; I) The published and widely used RT-qPCR assay for the E-gene [4, 5] combined with the SensiFAST^™^ Probe No-ROX One-Step Real-time PCR kit (Bioline®), and II) the commercial RealStar® SARS-CoV-2 RT-PCR kit 1.0 (Altona diagnostics, Hamburg, Germany). The RT-qPCR results (number of positives and Ct values) from the different approaches were compared to the RT-qPCR results from MagNA Pure purified samples. During this experiment the gravity of the limited supply for the MagNA Pure 96 system highlighted itself as our routine diagnostic laboratory became critically low on processing cartridges to the MagNA Pure 96 system, and we therefore had to switch to the QIAcube connect system (Qiagen, Hilden, Germany) to finish this study. The comparison of the simplified workflow to both NA purification systems is shown in Table 1, Table 2 and Figure 1, respectively.

**Table 1:**
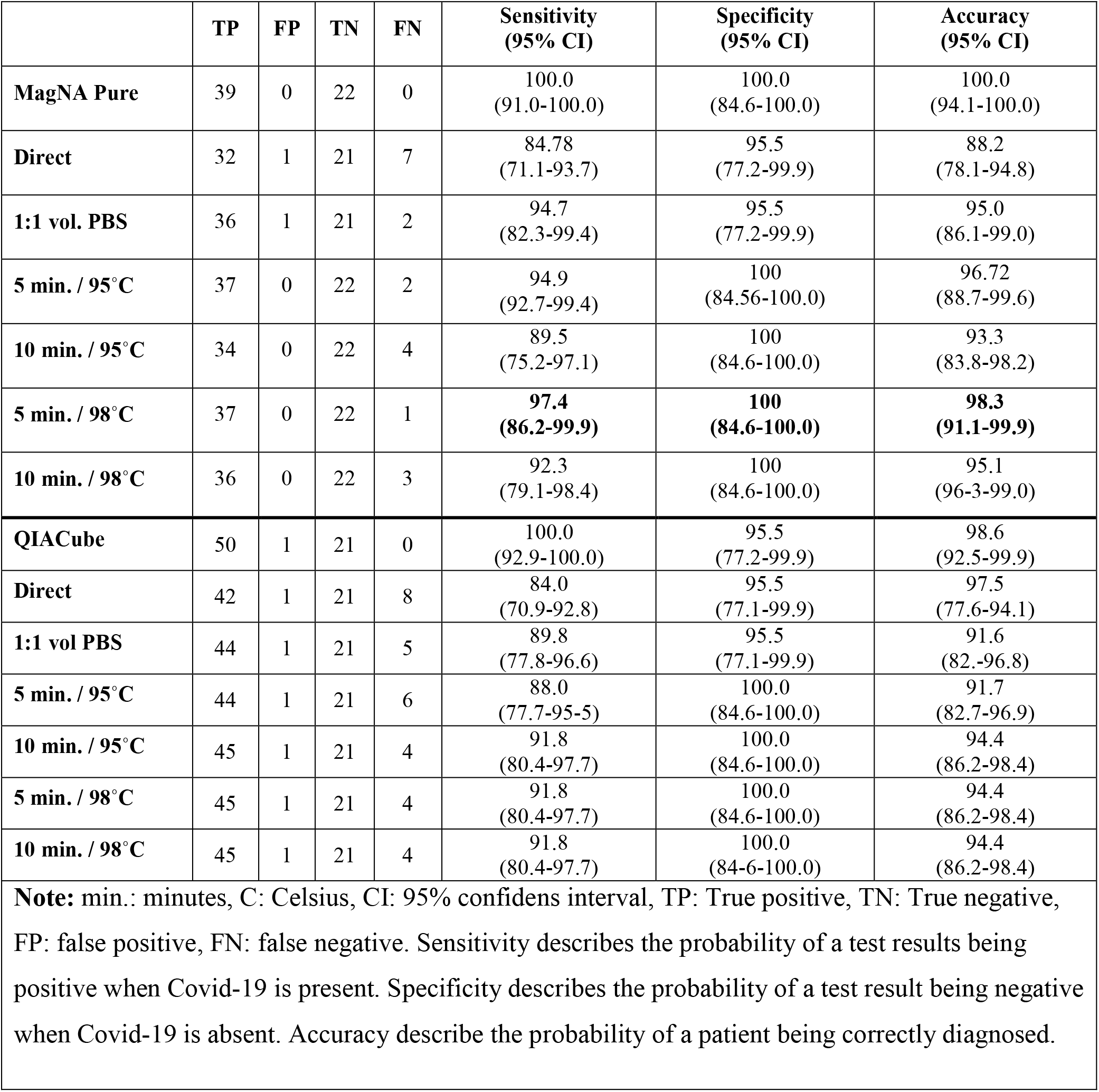
Analysis of clinical samples using the SensiFAST^™^ Probe No-ROX One-Step Real-time PCR kit.

|  | TP | FP | TN | FN | Sensitivity<br>(95% CI) | Specificity<br>(95% CI) | Accuracy<br>(95% CI) |
| --- | --- | --- | --- | --- | --- | --- | --- |
| **MagNA Pure** | 39 | 0 | 22 | 0 | 100.0<br>(91.0-100.0) | 100.0<br>(84.6-100.0) | 100.0<br>(94.1-100.0) |
| **Direct** | 32 | 1 | 21 | 7 | 84.78<br>(71.1-93.7) | 95.5<br>(77.2-99.9) | 88.2<br>(78.1-94.8) |
| **1:1 vol. PBS** | 36 | 1 | 21 | 2 | 94.7<br>(82.3-99.4) | 95.5<br>(77.2-99.9) | 95.0<br>(86.1-99.0) |
| **5 min. / 95°C** | 37 | 0 | 22 | 2 | 94.9<br>(92.7-99.4) | 100<br>(84.56-100.0) | 96.72<br>(88.7-99.6) |
| **10 min. / 95°C** | 34 | 0 | 22 | 4 | 89.5<br>(75.2-97.1) | 100<br>(84.6-100.0) | 93.3<br>(83.8-98.2) |
| **5 min. / 98°C** | 37 | 0 | 22 | 1 | **97.4**<br>**(86.2-99.9)** | **100**<br>**(84.6-100.0)** | **98.3**<br>**(91.1-99.9)** |
| **10 min. / 98°C** | 36 | 0 | 22 | 3 | 92.3<br>(79.1-98.4) | 100<br>(84.6-100.0) | 95.1<br>(96.3-99.0) |
| **QIAcube** | 50 | 1 | 21 | 0 | 100.0<br>(92.9-100.0) | 95.5<br>(77.2-99.9) | 98.6<br>(92.5-99.9) |
| **Direct** | 42 | 1 | 21 | 8 | 84.0<br>(70.9-92.8) | 95.5<br>(77.1-99.9) | 97.5<br>(77.6-94.1) |
| **1:1 vol PBS** | 44 | 1 | 21 | 5 | 89.8<br>(77.8-96.6) | 95.5<br>(77.1-99.9) | 91.6<br>(82.-96.8) |
| **5 min. / 95°C** | 44 | 1 | 21 | 6 | 88.0<br>(77.7-95.5) | 100.0<br>(84.6-100.0) | 91.7<br>(82.7-96.9) |
| **10 min. / 95°C** | 45 | 1 | 21 | 4 | 91.8<br>(80.4-97.7) | 100.0<br>(84.6-100.0) | 94.4<br>(86.2-98.4) |
| **5 min. / 98°C** | 45 | 1 | 21 | 4 | 91.8<br>(80.4-97.7) | 100.0<br>(84.6-100.0) | 94.4<br>(86.2-98.4) |
| **10 min. / 98°C** | 45 | 1 | 21 | 4 | 91.8<br>(80.4-97.7) | 100.0<br>(84.6-100.0) | 94.4<br>(86.2-98.4) |
| **Note:** min.: minutes, C: Celsius, CI: 95% confidens interval, TP: True positive, TN: True negative, FP: false positive, FN: false negative. Sensitivity describes the probability of a test results being positive when Covid-19 is present. Specificity describes the probability of a test result being negative when Covid-19 is absent. Accuracy describe the probability of a patient being correctly diagnosed. |  |  |  |  |  |  |  |

**Table 2:** Analysis of the median Ct values and interquartile range (IQR) for the detected and non-detected samples.

|  | **Median Ct value (IQR)** |  |
| --- | --- | --- |
|  | **Detected samples** | **Non-detected samples** |
| **MagNA Pure** | 28.7 (7.1) | 0.0 (0.0) |
| **Direct** | 32.0 (5.5) | 33.9 (2.5) |
| **1:1 vol. PBS** | 32.2 (6.7) | 35.1 (1.2) |
| **5 min. / 95°C** | 29.7 (6.9) | 33.0 (3.2) |
| **10 min. / 95°C** | 31.3 (6.4) | 32.7 (3.4) |
| **5 min. / 98°C** | 31.0 (7.2) | 29.8 (0.0) |
| **10 min. / 98°C** | 31.1 (6.3) | 34.7 (1.2) |
| **QIAcube** | 27.6 (8.6) | 0.0 (0.0) |
| **Direct** | 32.2 (6.1) | 34.6 (3.0) |
| **1:1 vol. PBS** | 31.0 (6.6) | 36.2 (10.7) |
| **5 min. / 95°C** | 30.4 (7.4) | 35.5 (3.7) |
| **10 min. / 95°C** | 30.6 (7.4) | 29.8 (9.5) |
| **5 min. / 98°C** | 30.5 (8.6) | 29.3 (8.2) |
| **10 min. / 98°C** | 30.3 (7.8) | 26.1 (8.4) |
| **Note: min.: minutes, C: Celsius, ct: cycle threshold, IQR: interquartile range** |  |  |

**Figure 1.**
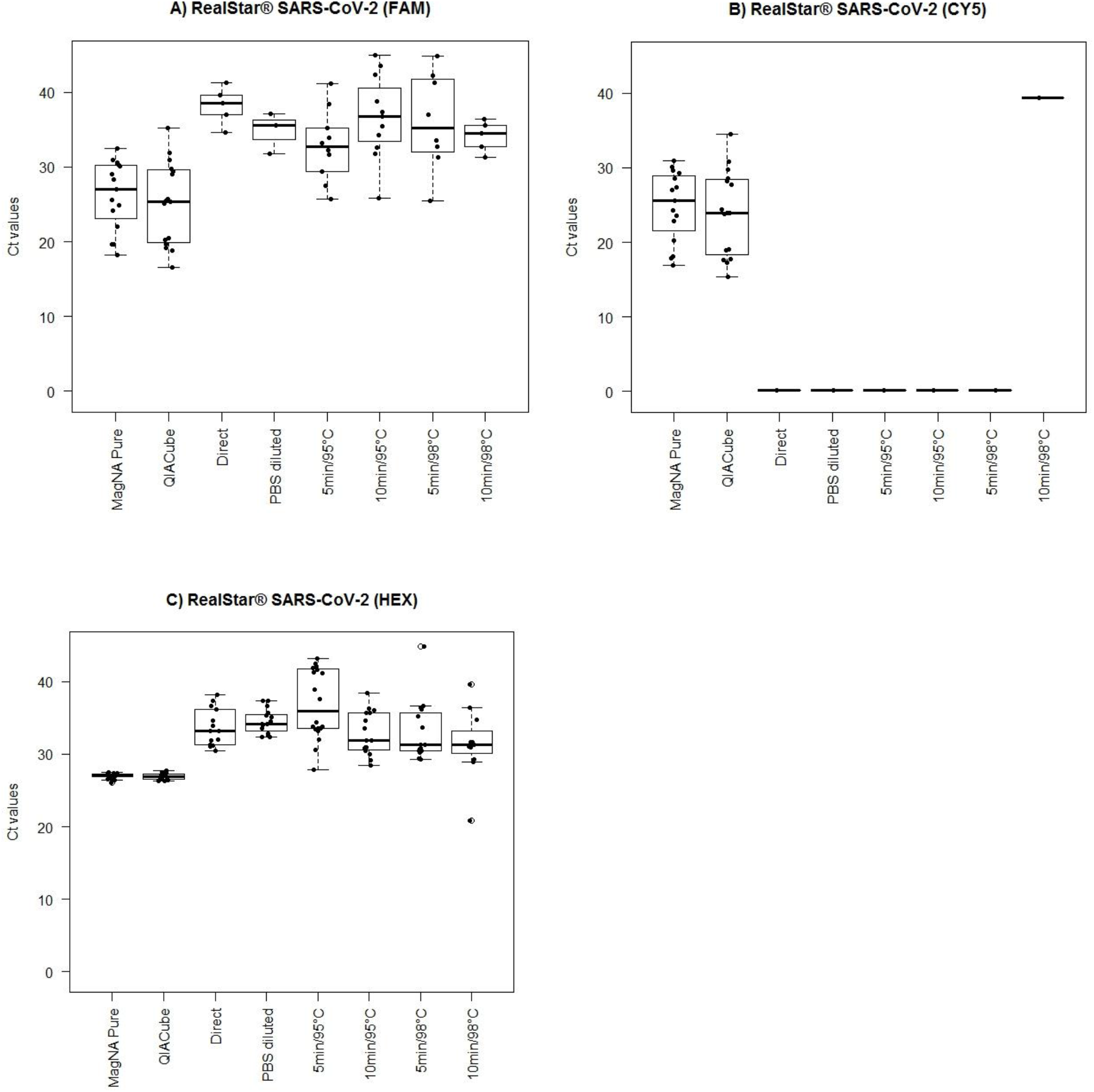
Median Ct values and interquartile range (IQR) for the RealStar® SARS-CoV-2 RT-qPCR assay. A) Detection of beta-coronavirus (FAM filter); B) Detection of SARS-CoV-2 (CY5 filter); C) Detection of the internal control (HEX filter). The RT-qPCR reactions were run on a MX3005P cycler (Stratagene) according to the manufactures instructions.

Analysis of SARS-CoV-2 positive and negative oropharyngeal swaps using the SensiFAST^™^ kit showed a 97.4% sensitivity, 100% specificity and 98.3% accuracy when samples were heat-processed for 5 min. at 98°C before the RT-qPCR reaction and compared to MagNA Pure purified samples (Table 1). False positive detection was observed for two of the non-heated samples (Ct=37) and for one QIACube purified sample (Ct=41.85), which could not be confirmed positive using either MagNA Pure purification, or any of the other simplified approaches. Overall, the simplified approach showed a lower sensitivity, specificity and accuracy when compared to QIACube purified samples than to MagNA Pure purified samples. Analysis of the median Ct values and interquartile range (IQR) for the detected and non-detected samples are shown in Table 2. In samples not detected there was a tendency toward high Ct values, but the pattern was not conclusive.

Analysis of the clinical samples using the RealStar® SARS-CoV-2 RT-PCR kit showed significant inhibition of the RT-qPCR reaction (Figure 1) except when the MagNA Pure and QIACube purified samples were used.

## Discussion

The newly emerged SARS-CoV-2 virus have challenged the global health system in every aspect including the ability to provide sufficient reagents for molecular diagnostic tests [2]. To overcome this shortening of supplies, computerised tomography (CT) scans of lungs have been used for diagnosis, with mixed results and at a risk of false-negatives especially during the early onsets of symptoms [6]. When a shortage in diagnostic kits in China happened, the Chinese health institutions diagnosed COVID-19 in a period in patients based on clinical symptoms alone, resulting in a major peak in the reported cases on February 12^th^ 2020 [7]. Because clinical symptoms for COVID-19 are sometimes non-specific (cough, mild fever, sore throat, fatigue), similar to other respiratory diseases or even absent despite infection [8, 9], molecular testing for SARS-CoV-2 [9] is necessary for a more correct diagnosis. In our diagnostic laboratory, purification of oropharyngeal swabs from patients is usually performed using the MagNA Pure NA purification system and diagnosis of COVID-19 is subsequently performed using the SensiFAST^™^ SARS-CoV-2 RT-qPCR assay. In this study we show that substitution of the MagNA Pure purification step with simple heating for 5 min. at 98°C will result in a sensitivity, specificity and accuracy of 97.4% (86.2-99.9), 100.0% (84.6-100.0) and 98.3% (95% CI=91.1-99.9), respectively, using the SensiFAST^™^ SARS-CoV-2 RT-qPCR assay (supplementary data). The SensiFAST^™^ SARS-CoV-2 RT-qPCR assay was superior in sensitivity to the RealStar® SARS-CoV-2 RT-qPCR kit 1.0, which was completely inhibited by the non-purified samples. We have not previously used the QIACube system to purify oropharyngeal swabs, and to our surprise QIACube purification of supposedly COVID-19 negative oropharyngeal swabs resulted in a weak but positive signal (Ct= 41.85) using the SensiFAST^™^ SARS-CoV-2 RT-qPCR assay. Moreover, this positive result could not be confirmed by any other NA purification method or SARS-CoV-2 specific RT-qPCR assay and therefore we cannot confirm if this patient sample is truly positive for COVID-19 using the QIACube purification system or false negative using the MagNA Pure system. The sensitivity, specificity and accuracy of the heating approach was lower when compared to samples confirmed positive by the QIACube. Due to this variation in sensitivity, specificity, and accuracy between different RT-qPCR assays, we recommend that all alternative RT-qPCR assay used together with the heat-processing workflow, should be validated before being implemented in clinical diagnostics.

Heating of the oropharyngeal swabs for 5 min. at 98°C followed by cooling for 2 min. at 4°C prior to a SARS-CoV-2 RT-qPCR reaction is not as sensitive or accurate as RT-qPCR reactions performed on purified samples. However, during a time where the spread of SARS-CoV-2 is so immense and molecular testing is critically challenged by the limited supplied of reagents for NA purification we may use alternative diagnostic methods. Simply heating of the samples could serve as an easy, fast and inexpensive alternative to chemical extraction kits, which would detect 97.4% of the COVID-19 positive patients with no false positives.

## Data Availability

The raw data (ct-values) generated in this study available on request

## Ethical statement

Exemption for review by the ethical committee system and informed consent was given by the Committee on Biomedical Research Ethics - Capital region in accordance with Danish law on assay development projects.

## Notes

**Conflict of interest:** None

### Competing Interest Statement

The authors have declared no competing interest.

### Funding Statement

No external funding was recieved

